# Plasma proteome reflects tissue damage and clinical manifestations in patients with inflammatory myopathies

**DOI:** 10.64898/2026.07.24.26358761

**Authors:** Yue-Bei Luo, Josefin Kenrick, Angeles S Galindo-Feria, Antonella Notarnicola, Ana Daniela Ulloa Navas, Irene Peralta-Garcia, Orane Demuynck, Andrea Kemp, Valerie Leclair, Karin Lodin, Eveline Van Gompel, Lara Dani, Fabricio Espinosa, Maryam Dastmalchi, Lina-Marcela Diaz-Gallo, Charlotta Preger, María Bueno Álvez, Mathias Uhlén, Peter Nilsson, Fredrik Edfors, Elisa Pin, Ingrid E Lundberg, Begum Horuluoglu

**Affiliations:** Division of Rheumatology, Department of Medicine, Solna, Karolinska Institutet, and Center for Molecular Medicine, Karolinska University Hospital, Sweden; Department of Neurology, Xiangya Hospital, Central South University, China; Department of Protein Science, SciLifeLab, KTH Royal Institute of Technology, Solna, Sweden; Department of Gastroenterology, Dermatology and Rheumatology, Theme Inflammation and Aging, Karolinska University Hospital, Stockholm, Sweden; Vall d’Hebron University Hospital, Barcelona, Spain; Division of Clinical Epidemiology, Department of Medicine, Solna, Karolinska Institutet, Stockholm, Sweden; Département de Médecine Interne et Immunologie Clinique, Centre Hospitalier Universitaire Lille, Centre de 1eference des Maladies Auto-Immunes Systémiques rares du Nord et Nord-Ouest, Lille, France; Department of Medicine, McGill University, Canada; Division of Rheumatology and Lady Davis Institute for Medical Research, Jewish General Hospital, Canada; Laboratory of Tissue Homeostasis and Disease, Skeletal Biology and Engineering Research Center, Department of Development and Regeneration, KU Leuven, Belgium

**Keywords:** inflammatory myopathy, myositis, proteomics, tissue damage, autoimmunity

## Abstract

**Objective:** Tissue-specific biomarkers that are reliable and associated with clinical manifestations of idiopathic inflammatory myopathy (IIM) are lacking. The blood circulation in individuals serves as a central conduit, allowing communication between tissues and facilitating clearance and recycling of tissue-derived proteins. Thus we applied a dual antibody-based proximity extension assay to profile the plasma proteome in subtypes of patients with IIM.

**Methods:** Plasma samples from 201 patients diagnosed with IIM at Karolinska University Hospital were analyzed using Olink. Clinical manifestations, disease activity measurements, and laboratory results were collected. First, we evaluated differential protein abundance across IIM subtypes. Then we calculated tissue scores for each disease subtype using tissue gene expression databases. Finally, interferon (IFN) and complement pathway scores were calculated.

**Results:** Plasma from patients with dermatomyositis was enriched for IFN signaling and skin associated proteins, anti-synthetase syndrome (ASyS) for lung associated proteins, immune-mediated necrotizing myopathy (IMNM) for muscle associated proteins, and inclusion body myositis for T cell response proteins. Patients with IMNM showed the highest tissue score for skeletal muscles, and ASyS for lung tissue score. Skeletal muscle scores correlated strongly with muscle disease activity visual analog scale scores, creatinine kinase levels, and muscle weakness score. Patients with interstitial lung disease along with and those with anti-Jo1 autoantibodies showed differential enrichment of surfactant proteins and IFN pathway activation.

**Conclusions:** Circulatory proteome is a promising tool to capture distinct tissue-specific responses that correlate with disease activity and tissue injury in patients with IIM.

## Introduction

Idiopathic inflammatory myopathies (IIM) comprise subtypes that affect organs such as skeletal muscles, skin, and lungs. Although clinical symptoms differ across clinical and serological subtypes, many shared pathological patterns are observed across organs, including inflammatory cell infiltration, microangiopathy, and fibrosis. These pathological processes reflect tissue injuries that manifest clinically. IIM are often categorized into five subgroups based on clinical manifestations and autoantibody profiles: dermatomyositis (DM), anti-synthetase syndrome (ASyS), immune mediated necrotizing myopathies (IMNM), inclusion body myositis (IBM), and polymyositis (PM). The most often screened myositis-specific autoantibodies (MSAs) include anti-Jo1, -PL7, -PL12, -OJ, -EJ, -SRP, -HMGCR, -Mi2, -MDA5, -NXP2, -SAE, and -TIF1γ autoantibodies. While MSAs are usually mutually exclusive, they can co-occur with myositis-associated autoantibodies (MAAs). One of the most common MAAs is anti-Ro52, which has been reported to be a marker of poor prognosis in some subsets of IIM [1, 2]. Autoantibodies are good diagnostic markers but are insufficient to capture the complexity of autoimmunity in IIM. Current blood biomarkers for clinical disease activity include creatinine kinase (CK) for muscle involvement and Krebs von den Lungen-6 (KL-6) for pulmonary fibrosis. However, they cannot be considered standalone surrogate for disease activity. Reliable, tissue-specific biomarkers associated with clinical manifestations and disease activity are needed but still largely lacking.

Developments in proteomics have greatly enhanced our capacity for biomarker identification. Modern platforms based on mass spectrometry or affinity-based proteomics allow broad and high-throughput coverage of the plasma proteome [3]. To date, several studies have used proteomics to investigate blood biomarkers in IIM [4–7]. These studies focused on identifying cytokines and a limited number of other immune molecules as biomarkers. One recent study using a protein panel of around 3000 molecules in juvenile DM found enriched lung proteins in patients positive for anti-MDA5 autoantibodies and increased extracellular matrix (ECM) modelling and angiogenesis in anti-NXP2 positive patients [8]. Still, an in-depth blood proteome analysis across all IIM subtypes and integrated with clinical phenotypes in adult patients is lacking.

In the present study, we applied a dual antibody-based proximity extension assay to profile plasma and evaluate protein differences across a well-characterized cohort of patients with IIM. We detected tissue-specific and clinically associated changes in the plasma proteome through correlation analysis with clinical variables. Our results provide insights into how circulating proteomic signatures may reflect organ involvement and disease activity in IIM.

## Methods

### Patient inclusion and sample acquisition

This study cohort consists of 201 patients with IIM from the Rheumatology clinic of Karolinska University Hospital that had plasma samples taken at time of diagnosis between 2002 and 2020. Patients fulfilled the 2017 EULAR/ACR classification criteria for idiopathic inflammatory myopathies [9]. We further subclassified patients into subgroups using the 2018 European Neuromuscular Center (ENMC) criteria for DM, Connors’ criteria for ASyS, the 2016 ENMC criteria for IMNM, and the 2011 ENMC criteria for IBM [10–13]. The PM subgroup was redefined as the PM patients remaining after patients with ASyS and IMNM were reclassified. MSAs and MAAs were profiled using the Euroimmun Myositis Profile-IgG (16 Ag) assay and chemiluminescence for HMGCR autoantibodies. The MSA status was used to subgroup patients by autoantibodies. All patients provided informed consent and this study was approved by the regional ethical committee in Stockholm region (2005/792-31/4).

### Protein detection

Plasma protein levels were measured using the Olink Explore 1536 panel as part of the Human Disease Blood Atlas program. Samples were fully randomized and normalized following standard plate protocol as described in Álvez [14]. Protein levels were reported in Normalized Protein eXpression (NPX). Eight proteins showing hook effect and a further 304 proteins with strong correlation with them were removed from downstream analysis [14].

### Protein differential abundance analyses

Differential abundance analysis (DAA) was performed using the limma algorithm, comparing one IIM subgroup against all other subgroups based on either diagnosis or autoantibody subgrouping. Multiple testing was corrected using the Benjamini-Hochberg method to control the false discovery rate (FDR). Proteins with FDR-adjusted p-values < 0.01 and |log2(fold change)| values > 0.13 were considered differentially abundant proteins (DAPs).

### Plasma protein correlations with clinical parameters

Clinical variables such as skin (including calcinosis, ulceration, mechanic’s hands), lung, muscle manifestations, Raynaud’s phenomenon, dysphagia, arthritis, and cancer status within one year of plasma sampling (N=193) were retrieved from prospectively collected data in the Swedish Myositis registry, SweMyoNet and patient records when needed. Laboratory variables include serum levels of CK, C reactive protein (CRP), and erythrocyte sedimentation rate (ESR). Disease activity variables included visual analogue scale (VAS) scores from the Myositis disease activity assessment tool (MDAAT, N=125), manual muscle test 8 scores (MMT8, N=115), and spirometry results (N=47) were collected from within 6 months of plasma collection, while laboratory test results were collected within 3 months of plasma collection (N=118-125). The TissueEnrich R package (v1.28.0) using the Human Protein Atlas was utilized to identify the tissue source of proteins that were significantly associated with clinical parameters [15].

### Tissue enrichment analysis

Gene expression from bulk RNA sequencing of 54 non-diseased tissues (v10) were obtained from the Adult Genotype-Tissue Expression portal (https://www.gtexportal.org/home/). The top 1000 enriched transcripts from 11 selected tissues were identified and used for the enrichment analysis for these tissues. Genes shared across all tissues were removed to increase tissue specificity. The derived RNA expression levels were used as a reference to define tissue enriched protein sets. Among these-tissue enriched proteins, 40 to 132 proteins from each tissue were present in the Olink panel (Supplementary Fig.1). For each tissue-enriched protein, a background protein not associated with the same tissue was randomly selected and its abundance level was subtracted from that of the tissue-specific protein for normalization purposes. The median of these protein-level differences was calculated for all proteins for the tissue. After 100 permutations, the final tissue score was defined as the median of these 100 median values.

### Pathway signature analysis

Scores for complement and interferon pathways were calculated using the mean NPX values of proteins listed for terms “complement cascade” from Reactome and “type I/II/III interferon (IFN) production”, “complement activation classical pathway” from Gene Ontology. The scores were calculated as the means of the proteins in the mentioned pathways. In addition, an IFN-inducible chemokine score was calculated based on the list curated from a previous study [16]. The alternative and lectin pathways of complement activation were not included in the analysis due to under-representation in our detection panel.

### Statistical analysis

Statistical analyses were performed in RStudio (v2025.05.0+496). Groups with a sample size smaller than five were removed from statistical analysis but were still plotted on relevant figures. Groupwise comparisons were performed using Kruskal-Wallis test followed by Dunn’s test. Differences in protein abundance in binary clinical variables were assessed using the Wilcoxon test with Bonferroni correction for multiple testing. Linear regression was used to evaluate association between proteins and continuous data. DAPs were defined as the ones with adjusted p-values < 0.05.

## Results

### Patient demographics and differential abundance of proteins in clinically and autoantibody defined subgroups

Anti-synthetase syndrome (ASyS, n=74) was the most common IIM subgroup, while PM was the least frequent (n=7, Table 1, Supplementary Fig.2). In terms of MSAs, anti-Jo1 autoantibody was the most frequent autoantibody (n=61). Moreover, 68 patients (34.7%) were positive for anti-Ro52 autoantibody as a single autoantibody or in combination with other MSAs. The overall female to male ratio was 1.6:1 and median age at sampling was 60 years (Table 1).

Differentially abundant proteins (DAPs) are listed in Supplementary Tables 1 and 2 (unadjusted and adjusted for age and sex, respectively). The age and sex adjustment did not significantly change the DAP profiles. As differences in age and sex may contribute to pathogenesis of IIM subtypes instead of acting purely as confounders, we chose not to adjust for age and sex in DAA [17, 18].

First, we analysed DAP profiles in clinically defined subgroups. Dermatomyositis (DM) was the clinical subgroup with the highest number of DAPs (n=104). The enriched DAPs included IFN pathway related (IFNL1, IFNγ receptor 2 [IFNGR2]), skin-associated (keratin 5/14 [KRT5/14]), ECM proteins (microfibril associated protein 5 [MFAP5], cathepsin O [CTSO]), and liver-associated proteins (alanine-glyoxylate aminotransferase [AGXT], sulfotransferase family 2A member 1 [SULT2A1], Fig.1A). Among the upregulated DAPs in the ASyS group, lung enriched proteins (surfactant protein D [SFTPD], mesothelin [MSLN]) and matrix proteins (myocilin [MYOC], Aggrecan [ACAN]) were detected (Fig.1A).

**Figure 1:**
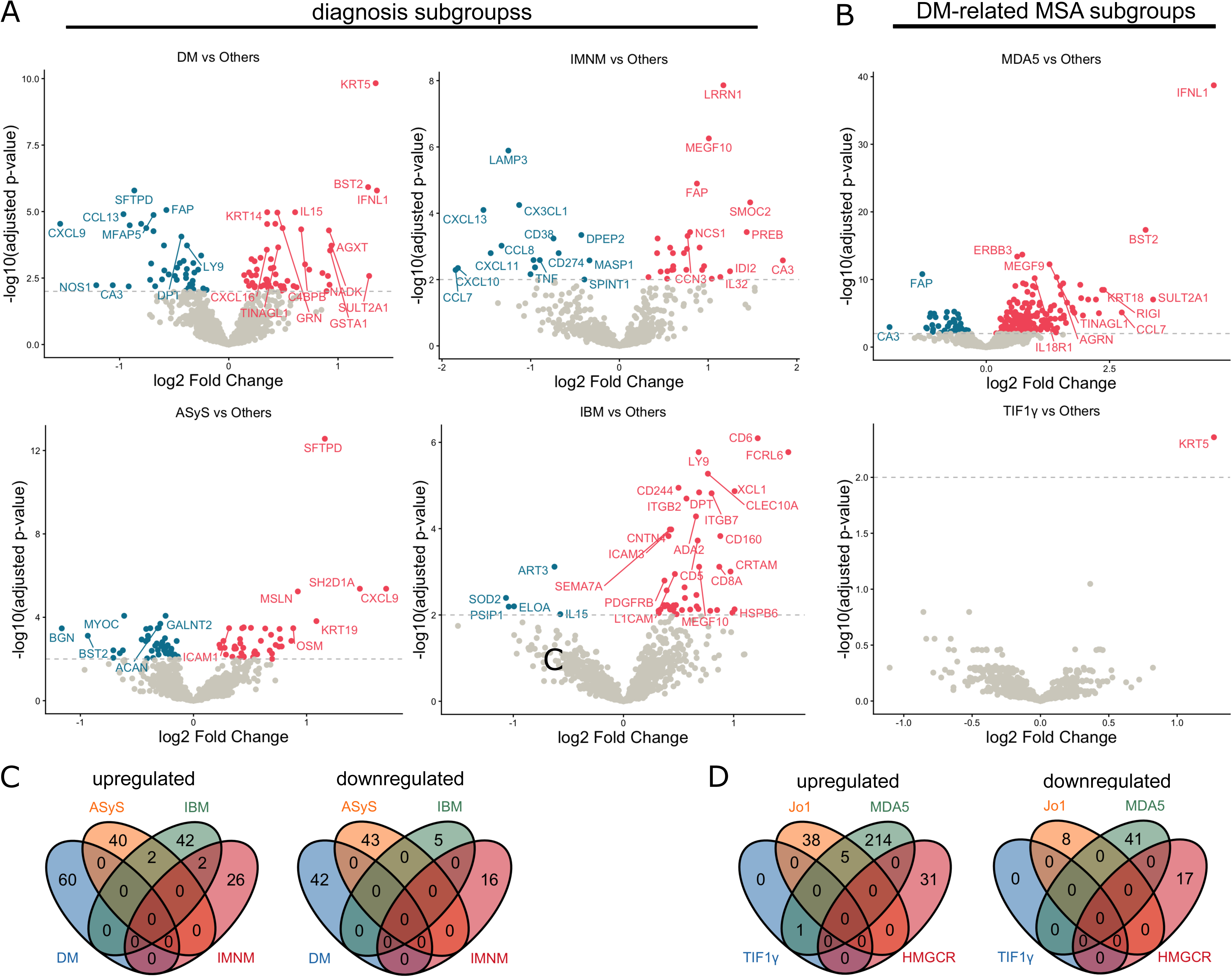
Differentially abundant plasma proteins across subgroups. Volcano plots showing DAPs in diagnosis subgroups of IIM (A) and autoantibody defined subgroups in DM (B). Venn diagrams showing overlapping DAPs across diagnosis (C) and autoantibody defined subgroups (D). Adjusted p-value cut off = 0.01, |log2FC| > 0.13, dots in red color = significantly increased, in blue = significantly reduced, in grey = non-significant.

In contrast, DAPs in the IMNM subgroup mainly consisted of muscle proteins (leucine rich repeat neuronal 1 [LRRN1], carbonic anhydrase 3 [CA3]), myogenesis (multiple EGF like domains 10 [MEGF10], neural cell adhesion molecule 1 [NCAM1]), and matrix proteins (fibroblast activation protein α [FAP], SPARC related modular calcium binding 2 [SMOC2]), Fig.1A).

The top enriched DAPs in patients with IBM were T cell response proteins (CD6, Fc receptor like 6 [FCRL6], lymphocyte antigen 9 [LY9]), followed by muscle proteins (heat shock protein family B member 6 [HSPB6], MEGF10) and ECM proteins (platelet derived growth factor receptor β [PDGFRB], scavenger receptor class F member 2 [SCARF2], Fig.1A). Interestingly, the IBM subgroup also demonstrated DAPs that are associated with nervous system development (contactin 1 [CNTN1], CNTN4, L1 cell adhesion molecule [L1CAM], neurotrophic receptor tyrosine kinase 2/3 [NTRK2/3]). There was no significant DAP detected between PM and the other subgroups.

Next we analysed DAPs in autoantibody defined subgroups. The anti-MDA5 autoantibody subgroup had the highest number of upregulated DAPs (n=263), whereas only one DAP was identified in the anti-Tif1γ positive group when compared to all other patients with IIM (Fig.1B). The anti-MDA5 subgroup, which is frequently associated with clinical lung involvement, was enriched in lung-specific proteins, such as surfactant protein A1 (SFTPA1) and KRT18. No DAP was detected in the seronegative DM group.

The ASyS group was dominated by anti-Jo1 autoantibody subgroup (83.6%). Therefore, the anti-Jo1 positive group was not different from the clinically defined ASyS subgroup analysis. Likewise, the anti-HMGCR autoantibody defined group accounted for 91.3% of the clinically defined IMNM, and the DAA results reflected the results from the comparison of IMNM against the other clinical subgroups (Supplementary Table 1). The anti-PL7 and anti-PL12 autoantibody subgroups did not show any DAPs, possibly due to small sample size.

### Tissue-enriched proteins associated with clinically distinct manifestations

Next, we explored the association between plasma proteins and laboratory variables and clinical manifestations, regardless of subgroups. Linear regression analysis revealed that 224 proteins were positively correlated with CK levels, which is a well-established biomarker for muscle injury (Fig.2A). Positively correlated proteins were most strongly enriched for skeletal muscle origin, with 3.02-fold more muscle-specific proteins than expected by chance (p=0.11), suggesting the skeletal muscle to be the primary source of these circulating proteins. In comparison, the systemic inflammatory marker CRP was associated with more heterogeneous tissue-enriched proteins such as FURIN (liver and salivary gland), TIMP metallopeptidase inhibitor 1 (TIMP1, ECM), and other pro-inflammatory molecules (IL6, colony stimulating factor 1 [CSF1], Fig.2B).

**Figure 2:**
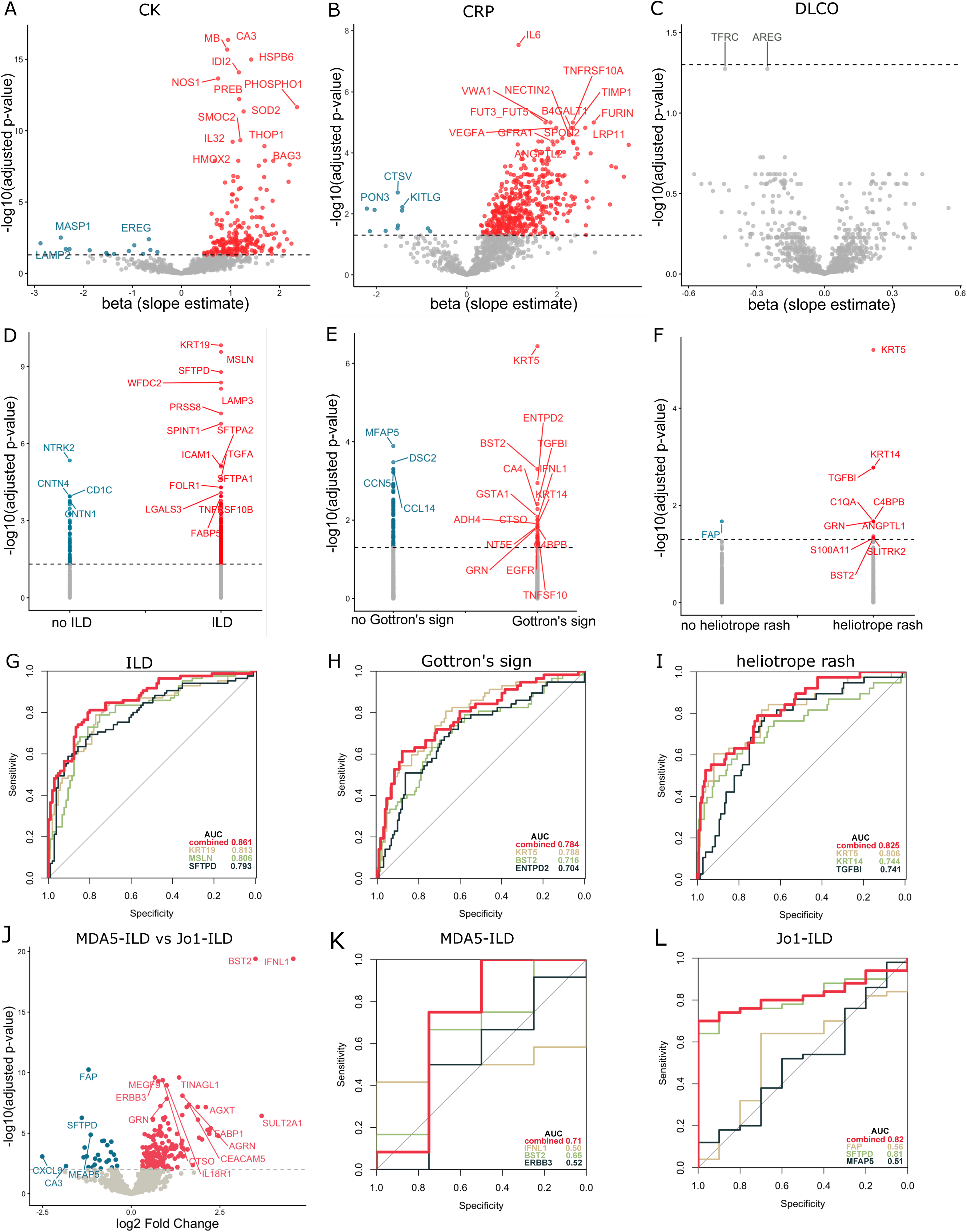
Association of differentially abundant proteins with clinical parameters. Volcano plots showing significant plasma proteins associated with CK (A), CRP (B), DLCO (C) levels, and ILD (D), Gottron’s sign (E), and heliotrope rash (F). ROC curves showing the performance of the top enriched DAPs in discriminating patients with ILD (KRT19, MSLN, SFTPD) (G), Gottron’s sign (KRT5, BST2, ENTPD2) (H) and heliotrope rash (KRT5, KRT14, TGFBI) (I) from those without. Volcano plot showing DAPs between MD5+ and Jo1+ patients with ILD (J). ROC curves showing the performance of the top enriched DAPs in discriminating patients with or without ILD in MDA5+ (IFNL1, BST2, ERBB3) (K), and Jo1+ (FAP, SFTPD, MFAP5) (L) groups. A-C, linear regression; D-F, Wilcoxon test with Bonferroni correction; J, limma algorithm. Adjusted p-value cut off = 0.01. A-F, dots in red color = significantly increased, in blue = significantly reduced, in grey = non-significant.

Interestingly, 234 proteins were significantly increased in patients with ILD compared to the others (Fig.2D, Supplementary Table 3). Tissue enrichment analysis demonstrated that the upregulated proteins exhibited a 1.82-fold enrichment for lung tissue, relative to the number expected under a random distribution (adjusted p-value=0.01). We compared anti-MDA5+ patients to anti-Jo1+ patients with ILD to identify DAPs associated with ILD in these two conditions. Patients with ILD in anti-MDA5+ autoantibody subgroup were enriched with type I/III IFN pathway proteins (such as IFNL1, BST2, RIG1), whereas patients with ILD in the anti-Jo1+ subgroup showed proteins more related to type II IFN pathway (such as CXCL9, CRTAM, KLRD1, Fig.2J-L, Supplementary Table 3). Interestingly, different surfactant proteins were also enriched between the two groups: patients with anti-MDA5 autoantibodies showed elevated SFTPA1 and SFTPA2, whereas those with anti-Jo1 autoantibodies had higher SFTPD levels (Fig.2J-L, Supplementary Table 3).

In patients with Gottron’s sign, skin was the most enriched tissue (3.98 times relative to expected, adjusted p-value=0.45, Fig.2E). Analysis using other categorical clinical parameters did not yield enough significant proteins for tissue enrichment analysis. The epidermal growth factor ligand amphiregulin (AREG) and the transferrin receptor (TFRC) showed a tendency towards negative association with predicted diffuse capacity of the lungs for carbon monoxide (DLCO) values (Fig.2C; p=0.05 for both). We selected the most differentially expressed proteins associated with ILD, Gottron’s sign, and heliotrope rash, and evaluated their ability to distinguish patients with or without these manifestations using receiver operating characteristic (ROC) curve analysis. These proteins demonstrated strong discriminatory performance, highlighting their potential as tissue damage biomarkers (Fig.2G-I). For ILD and heliotrope rash, combining the top DAPs further increased the AUC, reflecting improved performance in distinguishing patients with or without these symptoms.

The skin-associated proteins KRT5 and KRT14 were among the most enriched proteins in patients with DM that presented with skin rashes (Fig.2E, F, Supplementary Table 3). The IFN signaling and complement pathways related proteins were also highly enriched in these patients, suggesting IFN signaling and complement activation may be involved in the development of the skin rashes (Fig.2E, F). To further evaluate the overall activation of IFN signaling and complement pathways, we calculated and compared the pathway scores between patients with or without skin rashes as well as patients with ILD (Fig.3).

**Figure 3:**
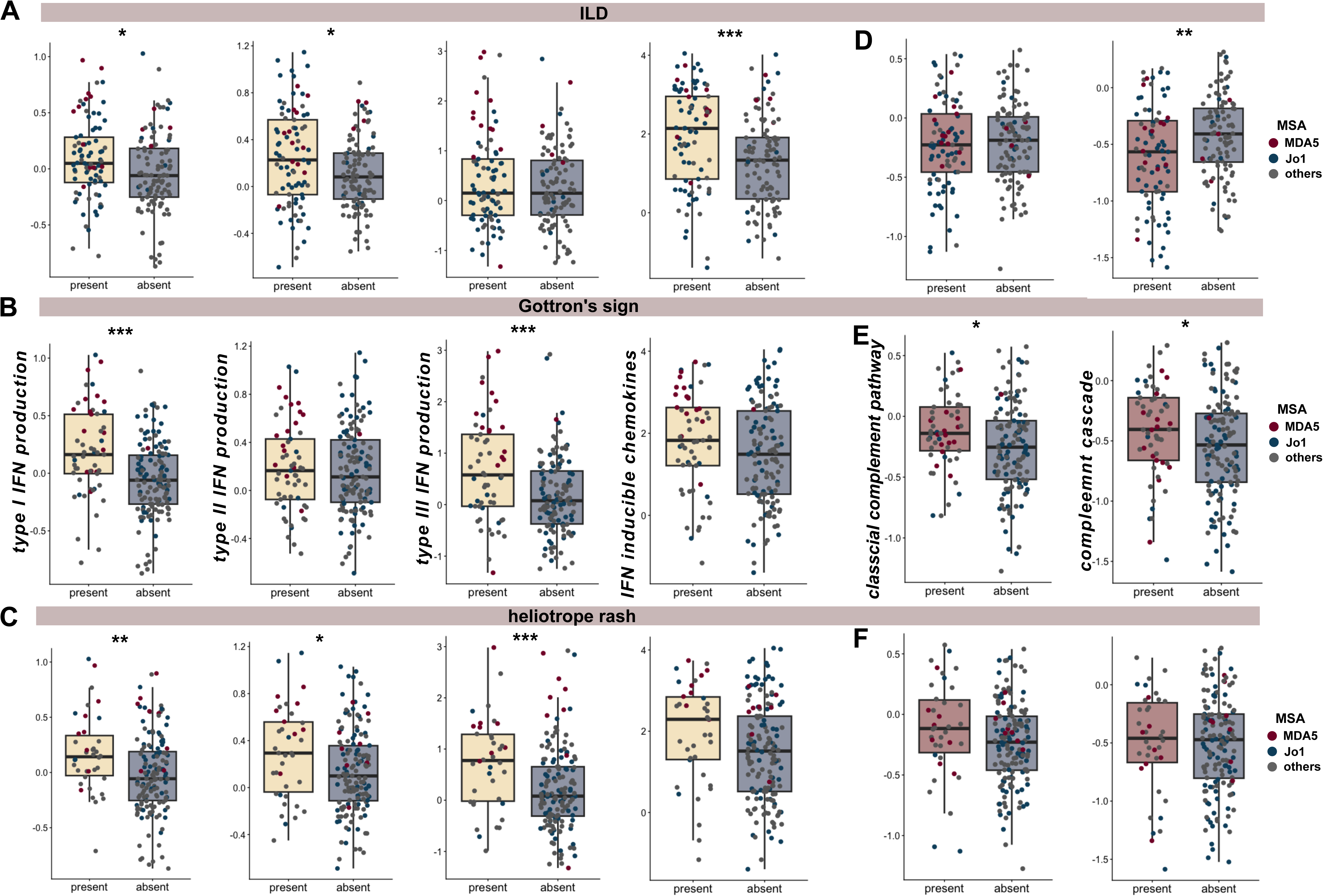
IFN and complement pathway scores among patients with distinct clinical manifestations. Boxplots demonstrating type I, II, III and IFN inducible (A-C) and complement pathway (D-F) scores in groups defined by ILD (upper panel), Gottron’s sign (middle panel) and heliotrope rash (bottom panel). Patients with anti-MDA5 and anti-Jo1 autoantibodies, two MSAs most commonly associated with lung and skin involvement, are indicated by different colors (red: anti-MDA5, blue: anti-Jo1, grey: others). Compared by Kruskal-Wallis test. * p < 0.05, ** p < 0.01, *** p < 0.001.

The scores for type I/II signaling and IFN induced chemokines in patients with ILD were higher compared to patients without ILD (p=0.01, 0.02, 1.7×10^-3^, Fig.3A). Gottron’s sign had higher scores for type I/III IFN signaling compared to patients without the manifestation (p=2.6×10^-5^, 5.6×10^-4^, Fig.3B). Patients with heliotrope rash had higher scores for production of all three types of IFN and IFN inducible chemokines (p=2.6×10^-3^, 0.02, 9.3×10^-4^, 0.02, Fig.3C). We also evaluated IFN signaling scores in ILD patients with anti-MDA5 or anti-Jo1 autoantibodies. Type I and III IFN production scores were higher in anti-MDA5 than in anti-Jo1 positive patients (p= 0.01, 3.18×10^-5^), while type II IFN scores were not different between the two subgroups (p=0.184).

The results regarding complement pathway were less consistent. Patients with ILD or heliotrope rash did not show any difference in classical complement pathway compared to their negative counterparts (p=0.59, 0.07), whereas patients with Gottron’s sign demonstrated activation of the classical complement pathway (p=0.01, Fig.3D, E). The complement cascade seemed to be enriched in patients with Gottron’s sign (p=0.04, Fig.3E, F), but was negatively enriched in patients with ILD (p=6.1×10^-3^, Fig.3D). This might be related to the relocation of complement related proteins into affected tissues from circulation.

We did not detect any DAP associated with pan-cancer diagnosis within 1 year of blood sampling or with individual cancer types. No DAP was identified for patients with Raynaud’s phenomenon, calcinosis, arthritis, mechanic’s hands, or dysphagia compared to patients without these clinical manifestations.

### Tissue scores differed between clinically and autoantibody defined subgroups

The presence of soluble tissue-related proteins in circulation that were associated with tissue manifestations prompted us to explore whether distinct tissue-specific signatures could be detected in the plasma of patients with IIM. For each tissue, a tissue score was calculated as the median expression levels of enriched proteins minus the background protein level.

Patients with IMNM had the highest skeletal muscle scores of all subgroups (p=3.8×10^-3^, Fig. 4A). The IBM group showed the lowest skin scores (p=0.03 for sun exposed skin, 2.5×10^-3^ for unexposed skin, Fig.4A). While the ASyS group demonstrated the highest lung scores of all clinically defined subgroups (p=8.1×10^-3^), the anti-MDA5 subgroup was the one with the highest lung score of the autoantibody defined subgroups (p=7.7×10^-4^, Fig.4B). The anti-MDA5 positive subgroup, surprisingly, also showed a high liver score compared to other subgroups (p=1.4×10^-5^, Fig.4B). The most enriched proteins for each tissue in our dataset are listed in Supplementary Table 4 and Fig.4C.

**Figure 4:**
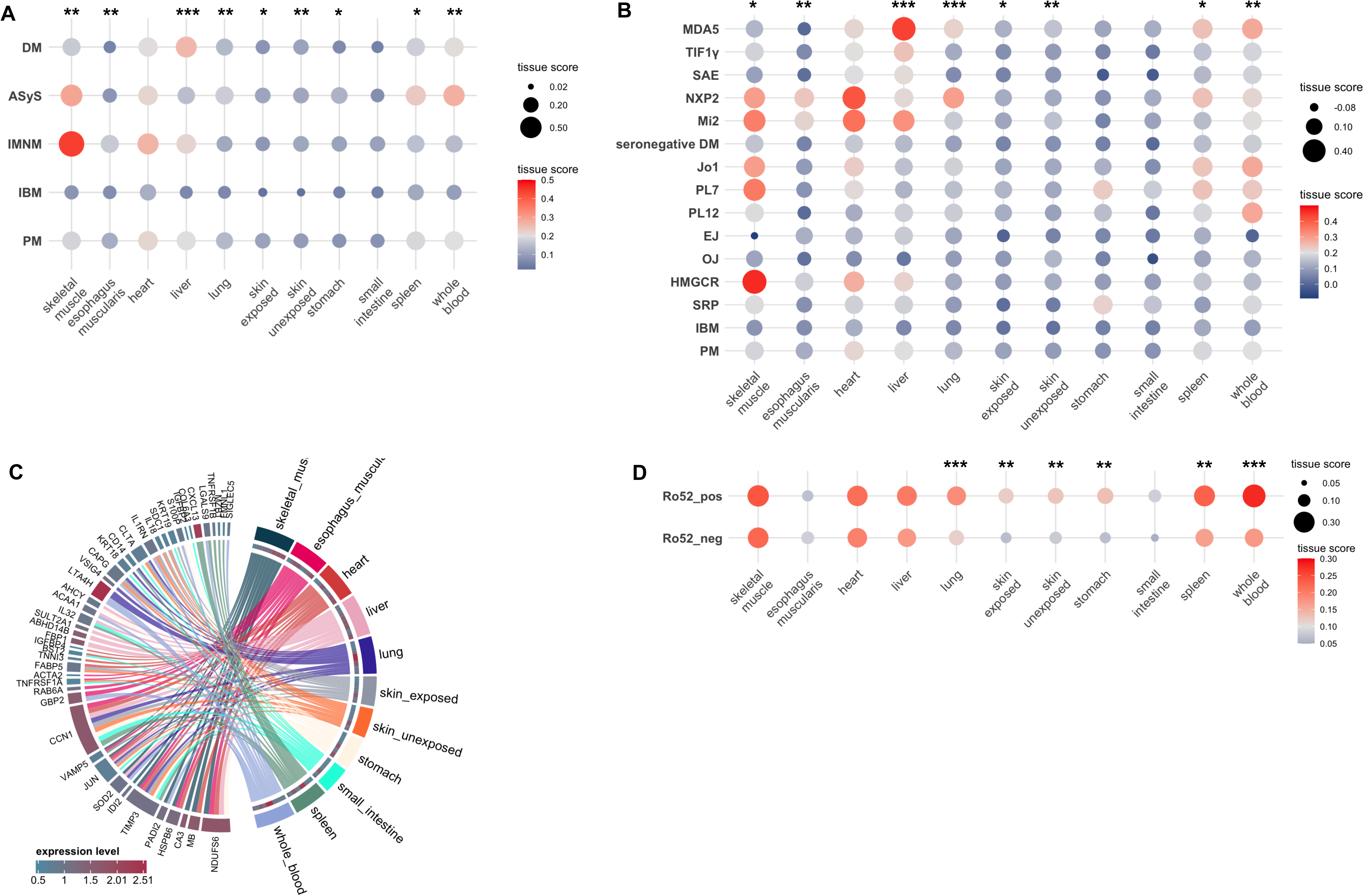
Tissue scores across diagnosis and antibody subgroups. Dot plots showing tissue scores in diagnosis subgroups (A) and autoantibody defined subgroups (B). Chord diagram showing the most enriched proteins in each tissue (C). Dot plot showing tissue scores in Ro52 autoantibody positive and negative subgroups (D). Kruskal-Wallis test with Bonferroni correction for A, B. Wilcoxon test with Bonferroni correction for D. Asterisk indicates significant overall difference across all groups. * p < 0.05, ** p < 0.01, *** p < 0.001.

In addition, patients with anti-Ro52 autoantibodies demonstrated higher skin and lung tissue scores than anti-Ro52 negative patients (p=1.3×10^-3^ for sun exposed skin, 4.2×10^-3^ for unexposed skin, 3.5×10^-4^ for lung, Fig.4D), but muscle scores were not different between anti-Ro52 positive and negative patients (p=0.63).

### Tissue scores correlated with disease activity and clinical symptoms

Next, we investigated tissue scores in relation to disease activity and laboratory results. The skeletal muscle scores in patients correlated strongly with both muscle disease activity (VAS muscle) and CK levels, as well as inversely with MMT8 (Fig. 5A). Lung tissue scores were strongly associated with VAS pulmonary and the systemic inflammatory markers CRP and ESR (Fig.5A). The levels of CRP showed correlations with a greater number of tissues in comparison to ESR, suggesting that it could be more pleiotropic than ESR.

**Figure 5:**
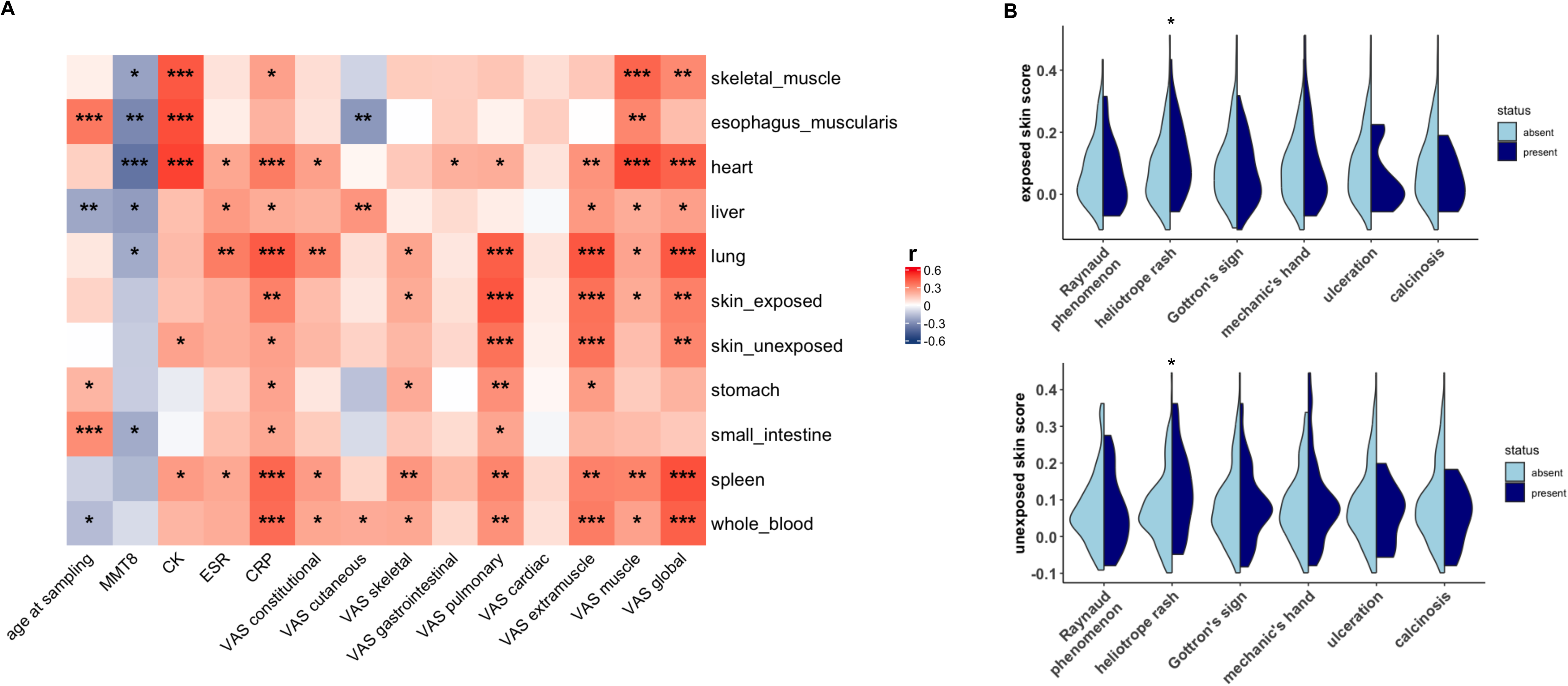
Tissue scores in association with disease activity and clinical symptoms. Heatmap demonstrating the correlation between tissue scores (y-axis) and clinical disease activity variables (A); violin plots showing the association of skin scores and skin-related symptoms (B). Kruskal-Wallis test with Bonferroni correction for A, and Wilcoxon test with Bonferroni correction for B. * p < 0.05, ** p < 0.01, *** p < 0.001. CK, creatine kinase; CRP, C reactive protein; ESR, erythrocyte sedimentation rate; MMT8, manual muscle testing 8; VAS, visual analogue scale.

Associations between clinical symptoms and tissue scores were also identified, particularly for skin and lung scores. Patients with heliotrope rash demonstrated higher skin scores than those without the rash (p=0.04 for sun exposed skin, 0.02 for unexposed skin scores, Fig.5B). Patients with ILD had significantly higher lung scores than those without ILD (p=6.3×10^-9^).

## Discussion

This study characterizes plasma proteins and the associations of plasma proteins with clinical features and disease activity in a large cohort of patients with IIM using dual antibody-based approach. First we identified DAPs associated with clinically and autoantibody defined IIM subgroups. We identified tissue-specific DAPs that corresponded to the preferentially affected organs. The subsequent tissue enrichment analysis based on calculated tissue scores confirmed that these altered proteins collectively reflect tissue involvement correlating with both disease subtypes and clinical involvement. Furthermore, our results support the notion that the blood proteome reflects the overall disease activity as well as distinct organ-specific activities. Together, these results highlight the potential of circulating proteomics as a comprehensive, system-level tool that might reflect organ involvement and to evaluate disease activity in IIM.

In systemic autoimmune diseases, immune responses occur across multiple sites in the body, including the circulation, lymphoid tissues, and target organs. In IIM, skeletal muscle, skin and lungs are the primary targets with occasional involvement of gastrointestinal tract, heart, and joints. Infiltrating immune cells within these tissues are potentially the culprits of tissue damage by disturbing cell metabolism, inducing cytolysis and cell death, compromising tissue regeneration, and promoting fibrosis [18, 19]. Our study indicated that tissue damage can be detected in the circulating proteins. This is consistent with a recent proteomic study of juvenile DM plasma, which also reported differential tissue involvement associated with the presence of different DM associated MSAs [8].

We calculated tissue scores derived from the relative expression levels of tissue-enriched proteins to evaluate how well circulating proteins represent tissue involvement. We chose tissue-enriched instead of tissue-specific proteins, since the numbers of the latter are small and vary significantly across tissues, which could introduce additional bias in tissue representation analysis using targeted proteomics. Consistent with clinical observations, the IMNM subgroup showed the highest skeletal muscle score in plasma samples, indicating severe muscle damage. The skeletal muscle score is mainly derived from muscle constituent proteins such as CA3, MB, and PADI2, which indicate active muscle necrosis, which is a typical myopathological feature in patients with IMNM. Importantly, the blood proteome not only captured tissue necrosis, but also other biological processes. The most differentially enriched proteins in IMNM include those related to muscle regeneration and ECM [20, 21]. It is well known that ECM remodeling is tightly regulated and closely involved during the dynamic muscle regenerative process [22, 23]. Thus, the blood proteome provides evidence of both active muscle necrosis and regeneration in patients with IMNM, which is markedly different from that in other IIM subgroups.

In contrast, the IBM subgroup demonstrated a different paradigm of circulating proteome disturbance. The top DAPs were enriched for proteins associated with T cell responses: LY9, which orchestrates T cell activation [24], and CD160, FCRL6, CD8A, and CD244 that are either directly involved in or regulate T cell cytotoxicity [25], reflecting typical myopathological features of patients with IBM. There were also several enriched T cell tissue retention markers (XCL1, integrin subunit beta 7 [ITGB7], cytotoxic and regulatory T cell molecule [CRTAM]) in IBM supporting the contribution of T cells in IBM pathogenesis. Previous work from our group has identified increased frequency of tissue resident memory T cells in IBM muscle identified by the expression of XCL1, XCL2, and CXCR6, suggesting that the muscle may be the source of the T cell related molecules in the circulation [26]. It has long been known that apart from myogenic changes, there are co-existing neurogenic changes in muscles indicated from electromyography of patients with IBM [13, 27]. However, nerve involvement has never been confirmed [28]. The presence of the nerve-related proteins such as GFAP and NEFL in plasma samples from patients with IBM may suggest enhanced muscle innervation to compensate for muscle atrophy that accumulates during the chronic disease progression or minimal nerve damage that remains undetectable by current electrophysiological techniques. We lean towards the former explanation, as classical nerve damage markers such as glial fibrillary acidic protein (GFAP) and neurofilament-light chain (NEFL) were not differentially abundant in IBM patients.

Our analysis also revealed high skin and lung protein abundance in the blood proteome of the DM and ASyS subgroups, respectively. The top DAPs KRT5 and KRT14 in patients with DM are markers of basal cells in airway epithelium and epidermis keratinocytes [29, 30]. A KRT5+KRT14+ subpopulation of basal cells with hyperplastic potential has been described in the lung of patients with idiopathic pulmonary fibrosis [30]. The abundance of these two proteins in DM may suggest either ongoing epithelium regeneration or ongoing skin damage. On the other hand, ASyS displayed a different set of lung- and skin-related DAPs compared to the DM subgroup. The top DAP, SFTPD, is a pulmonary surfactant glycoprotein and has been proposed as a biomarker for ILD and lung damage in other lung diseases [31–33]. In our study, SFTPD, along with MSLN expressed in pleural mesothelial cells and KRT19 in alveolar epithelial cells, was found to be strongly associated with ILD. Given that 84.9% of ASyS patients in this cohort had ILD, the observed protein differential abundance in ASyS is likely driven by or a result of the ILD.

Differences in protein abundance in patients with ILD were both distinct and nuanced when comparing the anti-MDA5 and anti-Jo1 autoantibody defined subgroups. For example, different types of IFN pathways characterize ILD in these two populations. This may indicate different pathophysiological mechanisms underlying the ILD development, with type I/III IFN contributing to ILD in anti-MDA5 and type II IFN to ILD in anti-Jo1 subgroup. As for the lung surfactant proteins, SFTPD was higher in anti-Jo1 positive ILD than in anti-MDA5 positive ILD, whereas SFTPA1 and SFTPA2 were higher in anti-MDA5 associated ILD. The former finding is in accordance with previous studies [34, 35]. Both SFTPA and SFTPD are involved in innate and adaptive immunity of the lung and can be pro- or anti-inflammatory in a context-dependent manner [36, 37]. Whether the differential abundance of the surfactant proteins reflects different ILD pathologies needs further study.

The blood proteome may also reveal subclinical organ involvement in IIM. We identified significantly elevated liver tissue scores in patients with anti-MDA5 autoantibodies. Indeed, liver dysfunction has recently been reported to be associated with anti-MDA5 autoantibody and a poor prognosis [38–40]. The evidence was largely restricted to laboratory tests such as γ-glutamyltransferase (GGT) and alkaline phosphatase (ALP). Imaging or pathology evidence of liver damage is sparse, with only case reports on nonspecific liver changes [41, 42]. The nature of liver damage thus remains unclear. The anti-MDA5 positive patients in this cohort demonstrated elevated GGT and ALP levels, but liver imaging did not show signs of liver involvement that warrants further investigation (data not shown). Our results might therefore suggest subclinical liver damage.

The present study provides evidence of higher lung and skin, but not muscle damage in the blood of anti-Ro52 autoantibody positive patients. Indeed, anti-Ro52 seropositivity has been associated with worse prognosis in patients with IIM, showing a particularly strong link to ILD, but not to muscle involvement [43–47]. There also seems to be a weak association between anti-Ro52 autoantibody and skin involvement [47, 48]. Our findings suggest that skin involvement is stronger in anti-Ro52 positive compared with anti-Ro52 negative patients. It remains to be elucidated whether damage in these tissues release TRIM21 into circulation and thereby may induce the production of anti-Ro52 autoantibody, or if the immune system targets TRIM21 in situ and thus causes tissue damage.

There are several limitations to this study. The tissue-sourced proteome in circulation is potentially affected by the different sizes, vascularization, and cell turnover rates of tissues. Although normalization and transformation of the tested protein levels minimize the range differences of tissue-specific proteins, they could still potentially affect the representation of tissue damage. This could probably explain the particularly good agreement of our proteome skeletal muscle scores and clinical muscle measurements such as VAS muscle scores, MMT8 and CK levels, while less strong correlations were observed between proteome skin scores and symptoms from the skin. Another major factor that could potentially bias tissue representation in the blood is the targeted protein detection panel. Although the present Olink panel enables detection of more than 1,000 proteins, untargeted proteome techniques such as mass spectrometry detects at least 3,000 proteins. Lastly, not all clinical data were collected in the registry at the time of sampling.

In conclusion, our results indicate that the blood proteome not only represents a complex repertoire reflecting systemic changes, but also, through careful dissection, could reveal distinct tissue-specific involvement. Moreover, these tissue responses on molecular level seem to correspond to tissue injury and disease activity in patients with IIM. Circulating proteomics thus offers a powerful approach to dissect tissue-level damage noninvasively in IIM.

## Data Availability

Boxplots of the NPX values can be found open access on The Blood Resource of the Human Protein Atlas
(https://www.proteinatlas.org/humanproteome/blood).

## Acknowledgement

W*e thank Dr Haris Babačić from Science for Life Laboratory and Department of* Oncology and Pathology, Karolinska Institutet for discussion about tissue enrichment analysis.

---

**Supplementary Figure 1:**
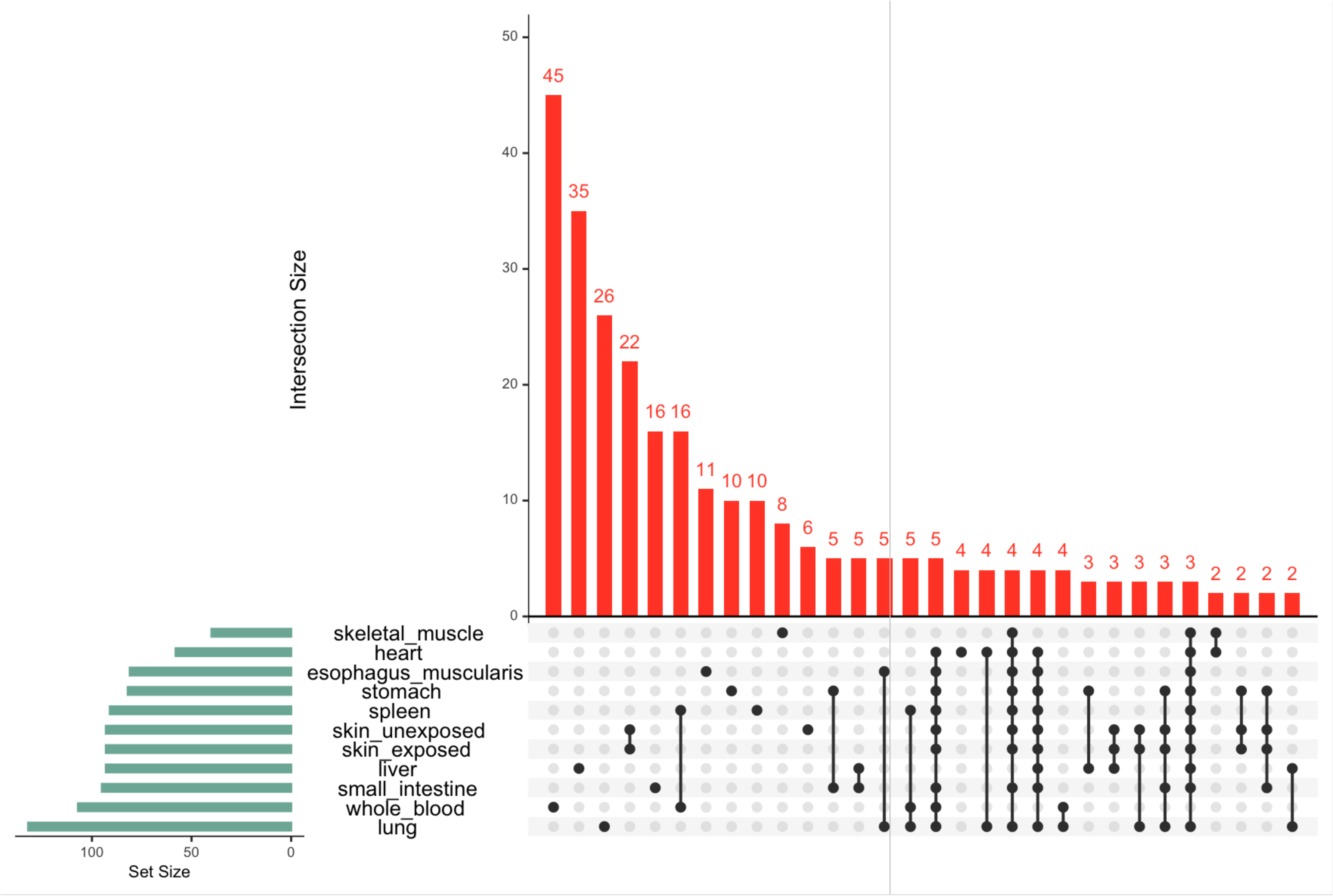
UpSet plot showing the proteins that overlap between defined tissue-specific protein sets among 11 selected tissues. Connecting dots show shared proteins across tissues, the numbers on the red columns show the total number of proteins shared or unique per tissue.

**Supplementary Figure 2:**
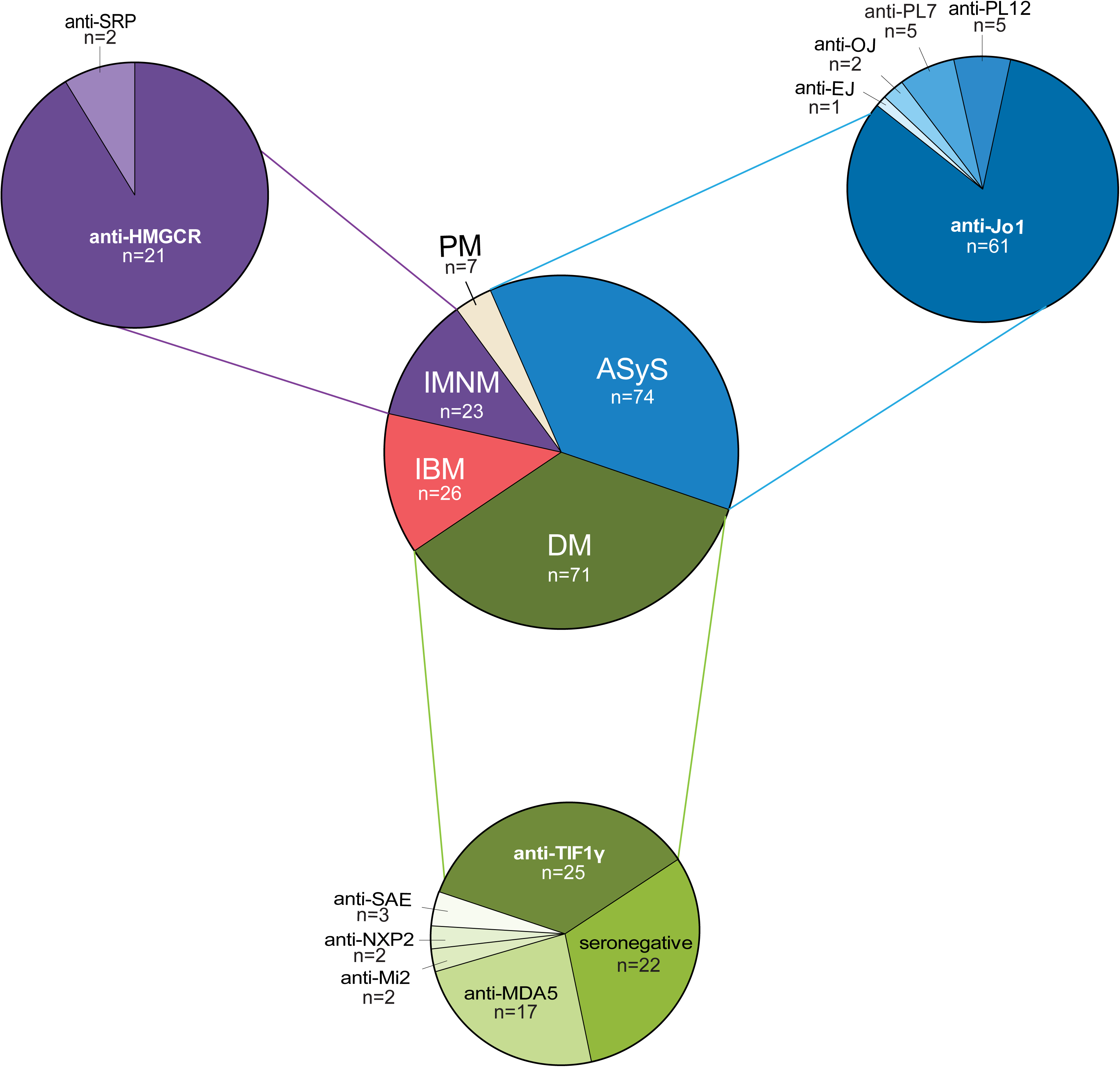
Patient composition based on diagnosis and autoantibody subgroups. Abbreviation: ASyS, anti-synthetase syndrome; DM, dermatomyositis; IBM, inclusion body myositis; IMMN, immune-mediated necrotizing myopathy; PM, polymyositis.

